# ‘Self-Management Intervention through Lifestyle Education for Kidney health’ (the SMILE-K study): protocol for a single-blind longitudinal randomised control trial with nested pilot study

**DOI:** 10.1101/2022.05.17.22275207

**Authors:** Courtney J. Lightfoot, Thomas J. Wilkinson, Thomas Yates, Melanie J Davies, Alice C. Smith

## Abstract

**Introduction:** Many people living with chronic kidney disease (CKD) are expected to self-manage their condition. Patient activation is the term given to describe the knowledge, skills, and confidence a person has in managing their own health and is closely related to the engagement in preventive health behaviors. Self-management interventions have the potential to improve remote disease management and health outcomes. We are testing an evidence- and theory-based digital self-management structured 10-week programme developed for CKD patients called ‘My Kidneys & Me’. The primary aim of the study (SMILE-K) is to assess the effect on patient activation levels.

**Methods and analysis:** A single-blind randomised control trial (RCT) with a nested pilot study will assess the feasibility of the intervention and study design before continuation to a full RCT. Individuals aged 18 years or older, with established CKD stage 3-4 (eGFR of 15-59 ml/min/1.73m^2^) will be recruited through both primary and secondary care pathways. Participants will be randomised into two groups: intervention group and control group. The primary outcome is the Patient Activation Measure (PAM-13). The full RCT will assess the effect of the programme on online self-reported outcomes which will be assessed at baseline, after 10-weeks, and then after 20-weeks in both groups. A total sample size of n=432 participants are required based on a 2:1 randomisation. A sub-study will measure physiological changes (e.g., muscle mass, physical function) and patient experience (qualitative semi-structured interviews).

**Ethics and dissemination:** This study was fully approved by the Research Ethics Committee-Leicester South on the 19/11/2020 (reference: 17/EM/0357). All participants are required to provide informed consent obtained online. The results are expected to be published in scientific journals and presented at clinical research conferences. This is protocol version 1.0 dated 27/01/2021.

**Trial registration number:** The study was prospectively registered as ISRCTN18314195 in December 2020.

## INTRODUCTION

Chronic kidney disease (CKD) is a long-term condition associated with high morbidity and premature mortality ^1^, and has an estimated UK prevalence ∼5–7%. In the UK, 70% of National Health Service (NHS) expenditure is spent on patients with long-term conditions such as CKD ^2^. With less than 1% of time spent in contact with healthcare professionals, many patients are expected to self-manage their condition ^2^. In CKD, the majority of people are managed in primary care rather than by kidney specialists ^3^. Long-term CKD management requires a high level of patient engagement, both in decision-making and in the implementation of care ^4^. For those with CKD, this encompasses a spectrum of behaviours ranging from adherence to medication and diet recommendations, maintaining physical activity, recognition and monitoring of risk factors (e.g., blood glucose and blood pressure), and self-adjustment of home-care routines ^5^. Self-management interventions aim to facilitate an individual’s ability to make appropriate lifestyle changes ^6^ and have shown beneficial impacts on various modifiable risk factors relevant to the progression of CKD (e.g. proteinuria, blood pressure, exercise capacity) ^6 7^. The COVID-19 global pandemic has presented unique challenges for people living with CKD and has further highlighted the need for and importance of self-management.

In order to implement self-management behaviours and participate in healthcare decisions, patients must have knowledge of their condition, and patient education is a crucial pathway to ensuring that individuals can be taught to engage in self-management tasks ^5^. Empirical studies have shown that patient education, including an understanding of CKD, is associated with better outcomes ^5^. Patient activation is the term given to describe the knowledge, skills, and confidence a person has in managing their own health ^8^ and is closely related to the engagement of preventive health behaviors ^4^. Studies have indicated that activated patients are more likely to attend screenings, check-ups and immunizations, as well as engage in healthy behaviors such as eating a balanced diet ^9 10^. Increased patient activation is associated with improved health outcomes in many long term conditions ^11 12^. In CKD lower patient activation is associated with worse cardiovascular disease risk profiles ^13^ and promoting patient activation in kidney disease care is increasingly being prioritized and has recently emerged as central to legislative policy in the United States ^14^ and UK ^15^. In the UK, National Institute for Health and Care Excellence (NICE) clinical guidance recommends that informational and educational programmes are offered to those with CKD, including information regarding what people can do to manage and influence their own condition. Interventions to increase patient activation are likely to improve self-management behaviour, and consequently this may be a suitable outcome for self-management-based interventions.

A range of barriers have prevented widespread implementation of comprehensive education for people with CKD and a recent systematic review of self-management interventions in CKD found many interventions lack patient engagement in their design, and the majority of interventions do not apply behavioural change theory to inform their development ^7^. As such there remains a need for better and innovative self-management interventions for those with CKD ^3 7^. Digital self-management interventions have the potential to improve remote disease management and health outcomes ^16^ and are increasingly becoming integrated into self-management to improve behaviour. The COVID-19 pandemic has also presented those with long-term conditions and their healthcare teams an opportunity to innovate and move towards an increasingly digitalised care, with particular emphasis on supporting patients from their own homes.

Developed in the UK, the structured DESMOND diabetes self-management group education programme has recently been shown to improve patient activation in individuals living with diabetes ^17^. Based on the same principles, MyDESMOND is a global digital programme to provide ongoing support and guidance ^18 19^. Using the MyDESMOND platform, we developed an evidence- and theory-based digital self-management programme for CKD patients called ‘My Kidneys & Me’. The programme was developed in conjunction with patients and their families, key stakeholders, and a wide range of healthcare professionals including nephrologists, psychologists, physiotherapists, dieticians, exercise scientists, and pharmacists. The programme was developed using ‘Intervention Mapping’, a six-step framework used to guide behaviour change interventions and health education development ^20^. A full and detailed description of the development of ‘My Kidneys & Me’ can be found in Lightfoot et al. ^21^.

### Objectives and hypotheses

The primary aim of the proposed ‘SMILE-K’ research study is to assess the effect of a structured online self-management programme - ‘My Kidneys & Me’ - on patient activation levels and subsequent self-management behaviour. Further objectives include assessing the feasibility of using such an intervention in this population and exploring patient experience of the programme itself. We hypothesise that access to ‘My Kidneys & Me’ will increase patient activation, compared to usual care, in people living with CKD.

## METHODS AND ANALYSIS

This protocol adheres to the SPIRIT (Standard Protocol Items: Recommendations for Interventional Trials) reporting recommendations ^22^ (Supplementary material S1).

### Study design overview

A single-blind randomised control trial (RCT) with a nested pilot study will be used to assess the effect of the ‘My Kidneys & Me’ programme. An initial nested pilot study will assess the feasibility of the intervention and study design before continuation to a full RCT is considered. Continuation to the full RCT will be based on a pre-defined ‘stop/go’ criteria assessed after n=60 participants have been recruited. The study intervention period will last 20-weeks with outcome measures assessed at baseline (pre-intervention), week-10 (post-intervention), and week-20 (follow-up). A flow diagram showing the participant flow through the study can be found in **Figure 1**.

**Figure 1.**
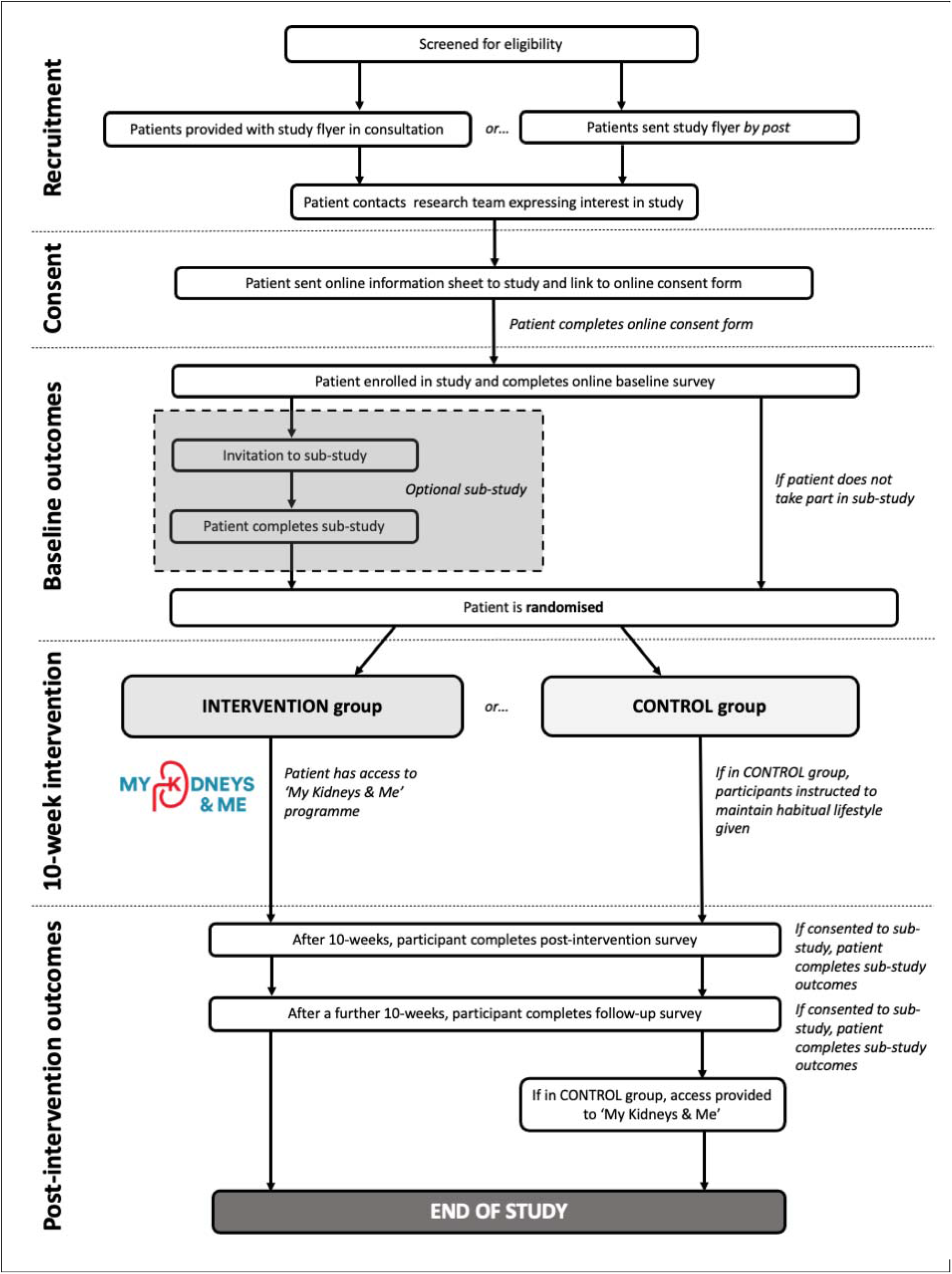
Study flow diagram

An optional sub-study will occur in parallel with the main study. This will consist of additional objective assessments of body composition, physical activity, and physical function to the main study outcome protocol, as well as qualitative interviews with participants to discuss their expectations and experiences of the intervention and study protocol.

### Eligibility criteria

Individuals aged 18 years or older, with established CKD stage 3-4 (eGFR of 15-59 ml/min/1.73m^2^) according to the NICE guidelines will be included. Those requiring any form of kidney replacement therapy (i.e., any modality of dialysis, or transplantation) or with insufficient command of English or any other precluding factors that prevent ability to give informed consent or comply with protocol will be excluded. For the sub-study, in addition, participants will be excluded if they are pregnant and/or if any element of protocol considered by own clinician or General Practitioner (GP) is contraindicated and/or the individual is deemed unfit due to physical impairment, significant co-morbidity, or other reason (e.g., unstable hypertension, arrhythmia, myocardial infarction <6 months, unstable angina, uncontrolled diabetes mellitus, advanced cerebral or vascular disease).

### Recruitment

Participants will be recruited through both primary and secondary care pathways across multiple sites in England. Primary care practices will identify eligible patients and provide them with an introduction flyer and study invitation letter, either during a consultation or via the post. If recruited from secondary care, the flyer will be provided during the patient’s routine outpatient clinic visit by the nephrologist. Alternatively, patients may be recruited by postal means. Interested participants are requested to contact the research team by email who will then respond with further detailed information and an online consent form. When participants have consented, they are sent a link to complete the online outcome measures. In eligible patients, further information regarding the sub-study is provided. Once the baseline outcome measures have been completed, participants are randomised and provided access to the programme if appropriate. Access will be a secure link that ask participants to register and create a unique username and password. Once created, patients can then access ‘My Kidneys & Me’ (dashboard shown in **Figure 2**).

**Figure 2.**
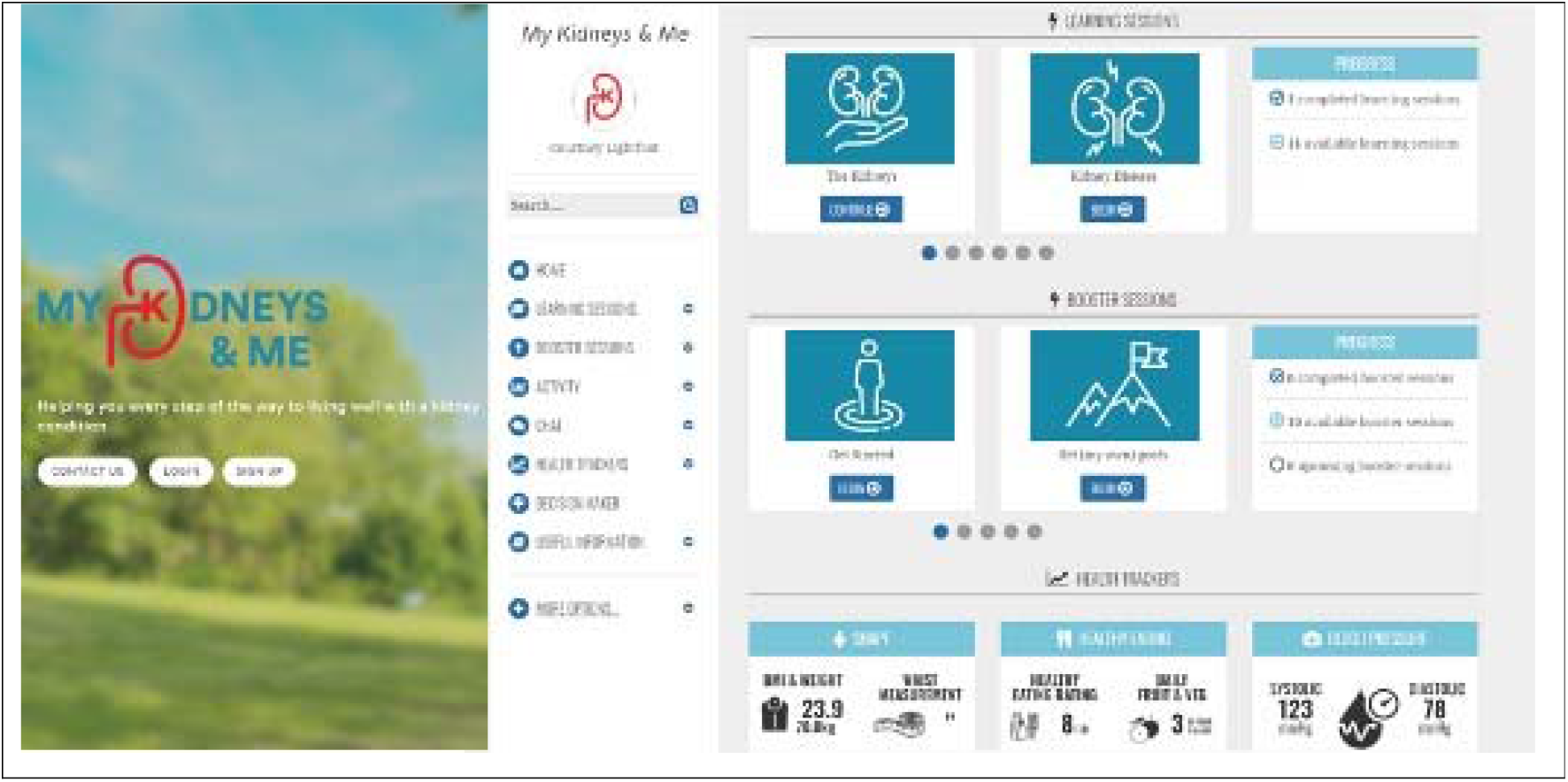
‘My Kidneys & Me’ dashboard page

### Randomisation

Participants will be randomised into two groups: intervention group and control group. Randomisation will be performed by the research team in a single-blind fashion. Participants will be stratified based on age (≤63, >63 years) to ensure comparatively equal representative age characteristics in both groups. These values are based on the median age attained from preliminary unpublished data from two ongoing observational studies in non-dialysis CKD patients by our group. The control group will not be provided with the intervention during the study and will be asked to maintain their habitual lifestyle activities. The control group will be provided with the intervention upon study completion. The intervention group will be provided access to the online ‘My Kidneys & Me’ intervention.

### Intervention

#### ‘My Kidneys & Me’

The ‘My Kidneys & Me’ programme forms part of the award-winning and quality assured MyDESMOND e-learning platform ^21^. The educational sessions provide information about the kidneys, CKD, its treatment, and the different ways to self-manage and is written to accommodate for those with low patient activation levels. ‘How to’ booster sessions are interactive educational sessions, which provide instructions on how to perform the self-management behaviours and are released weekly. A summary of these sessions is found in **Table 1**.

**Table 1.**
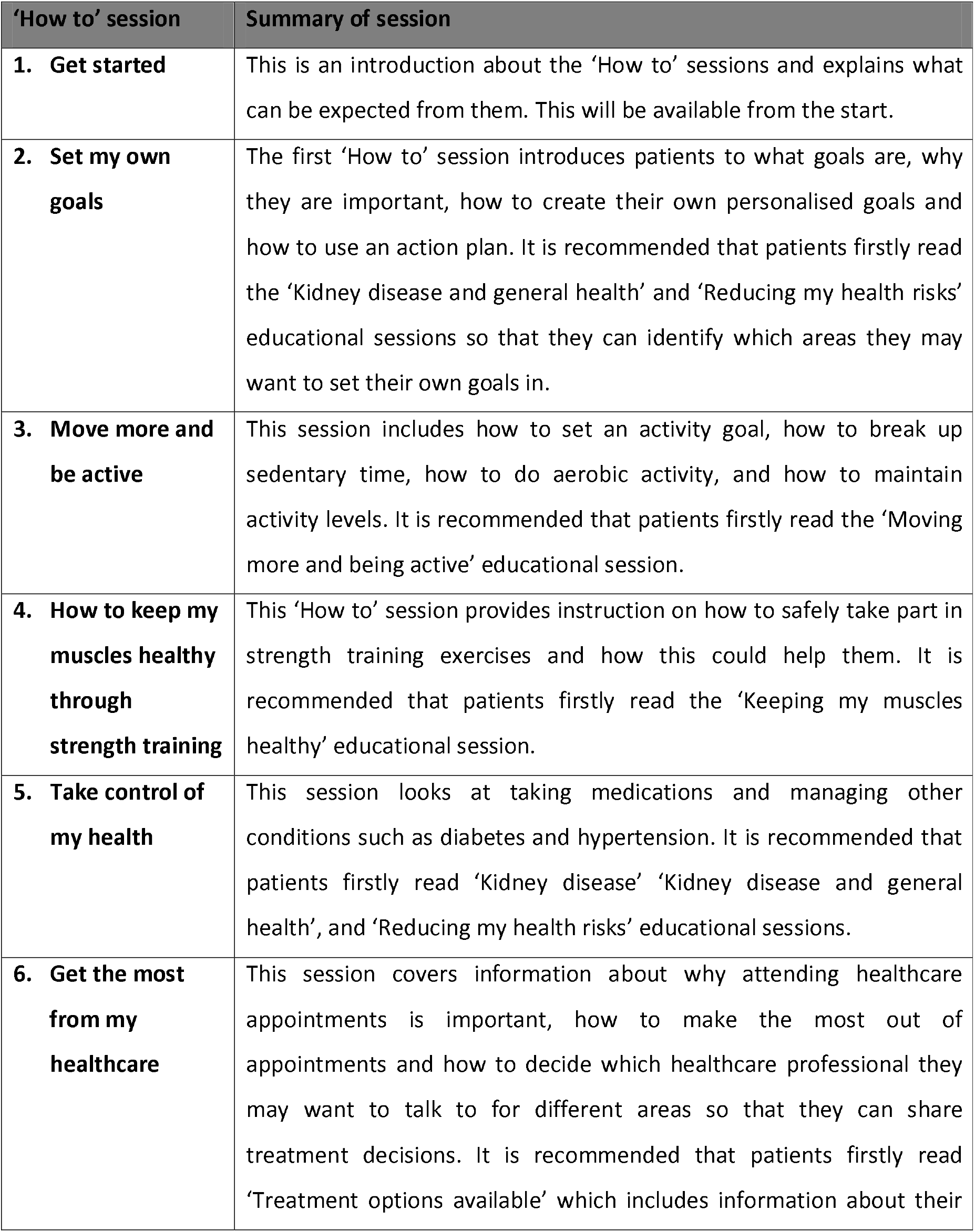
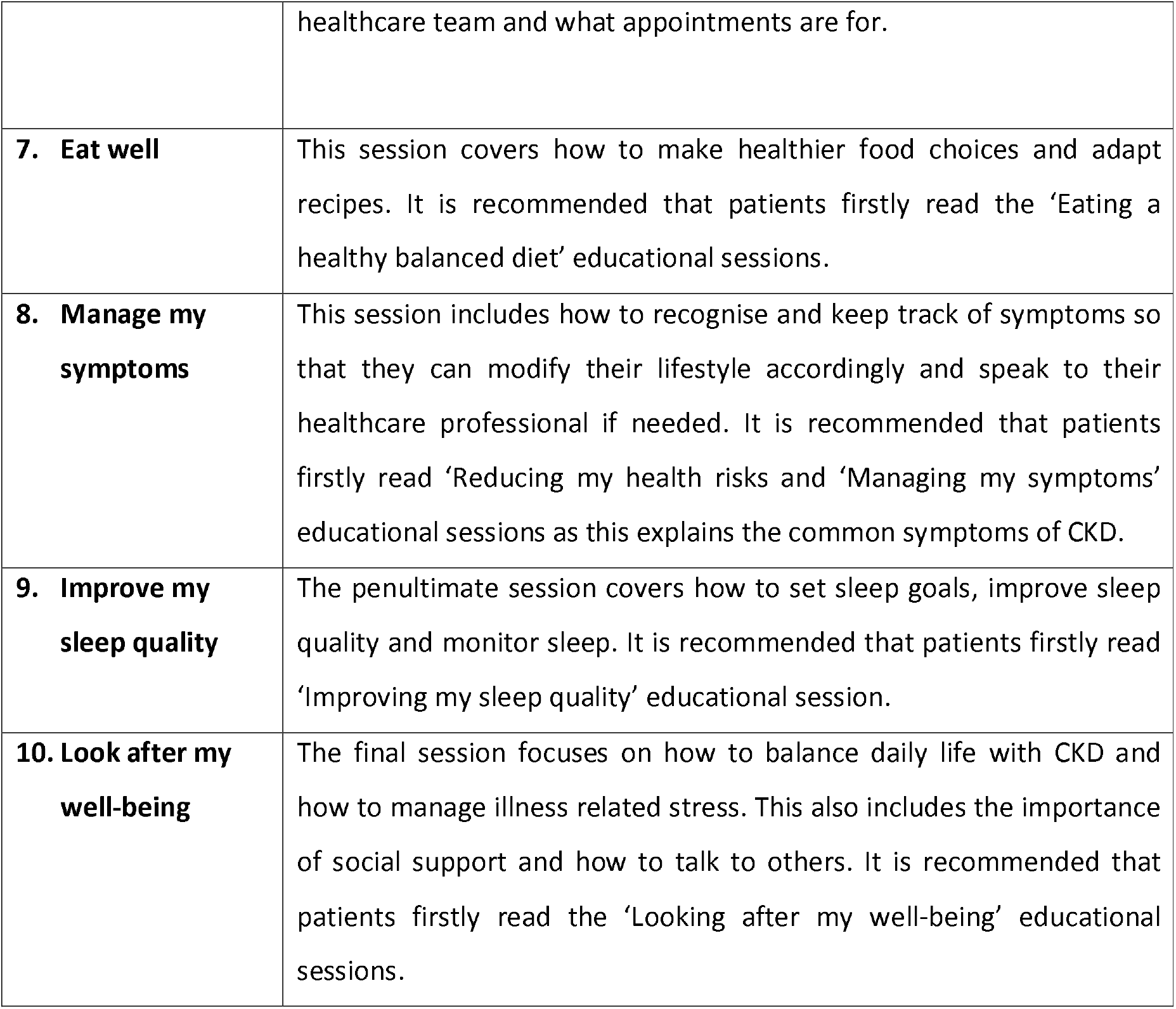
Summary of the ‘How to’ booster sessions.

| **‘How to’ session** | **Summary of session** |
| --- | --- |
| **1. Get started** | This is an introduction about the ‘How to’ sessions and explains what can be expected from them. This will be available from the start. |
| **2. Set my own goals** | The first ‘How to’ session introduces patients to what goals are, why they are important, how to create their own personalised goals and how to use an action plan. It is recommended that patients firstly read the ‘Kidney disease and general health’ and ‘Reducing my health risks’ educational sessions so that they can identify which areas they may want to set their own goals in. |
| **3. Move more and be active** | This session includes how to set an activity goal, how to break up sedentary time, how to do aerobic activity, and how to maintain activity levels. It is recommended that patients firstly read the ‘Moving more and being active’ educational session. |
| **4. How to keep my muscles healthy through strength training** | This ‘How to’ session provides instruction on how to safely take part in strength training exercises and how this could help them. It is recommended that patients firstly read the ‘Keeping my muscles healthy’ educational session. |
| **5. Take control of my health** | This session looks at taking medications and managing other conditions such as diabetes and hypertension. It is recommended that patients firstly read ‘Kidney disease’ ‘Kidney disease and general health’, and ‘Reducing my health risks’ educational sessions. |
| **6. Get the most from my healthcare** | This session covers information about why attending healthcare appointments is important, how to make the most out of appointments and how to decide which healthcare professional they may want to talk to for different areas so that they can share treatment decisions. It is recommended that patients firstly read ‘Treatment options available’ which includes information about their |
|  | healthcare team and what appointments are for. |
| **7. Eat well** | This session covers how to make healthier food choices and adapt recipes. It is recommended that patients firstly read the 'Eating a healthy balanced diet' educational sessions. |
| **8. Manage my symptoms** | This session includes how to recognise and keep track of symptoms so that they can modify their lifestyle accordingly and speak to their healthcare professional if needed. It is recommended that patients firstly read 'Reducing my health risks and 'Managing my symptoms' educational sessions as this explains the common symptoms of CKD. |
| **9. Improve my sleep quality** | The penultimate session covers how to set sleep goals, improve sleep quality and monitor sleep. It is recommended that patients firstly read 'Improving my sleep quality' educational session. |
| **10. Look after my well-being** | The final session focuses on how to balance daily life with CKD and how to manage illness related stress. This also includes the importance of social support and how to talk to others. It is recommended that patients firstly read the 'Looking after my well-being' educational sessions. |

The health trackers feature allows patients to self-monitor different aspects of their health that are involved in the self-management of the condition: body weight and measurements; fruit and vegetable intake; symptoms; smoking; cholesterol; and blood pressure. These allow patients to manually update their trackers and see their progress over time through graphs and charts. Similarly, the ‘Activity’ feature allows patients to track their physical activity, although this is broken down into different forms of activity. Patients can track how many steps they have walked and for how many minutes and can challenge others or invite up to five of their own friends and family to join these challenges. The patients can track this by either synchronizing the programme with their activity tracker (e.g., Fitbit) or they can enter their steps manually. They can also use a bespoke tracker created for ‘My Kidneys & Me’ to record their strength training progress and resources (e.g., instructional videos) will be available online. The Decision maker feature is a tool to help patients create and monitor their own health related goals, as this works through a series of questions to help patients identify which goals are most important to them and how they can achieve these by overcoming identified barriers.

### Outcome measures

#### Nested feasibility outcomes

Before progression to the full RCT, we will assess feasibility outcomes on the first n=60 participants recruited. Based on these outcomes, a decision will be made to continue with the trial based on pre-specified progression criteria. These progression criteria will be developed with input from stakeholders in the study, researchers, and patients. The following feasibility outcomes will be assessed:

- *Recruitment rate:* The number of eligible patients and number consented will be recorded. Monthly recruitment rate and the time taken to recruit 25%, 50%, 75%, and 100% patients will be recorded.
- *Acceptability of randomisation and assessment procedure:* Acceptability of randomisation (and stratification variables) and procedures will be determined by comparison of randomized group characteristics, and by measuring loss to follow-up and by exploring patient’ views about their participation in the research.
- *Programme usage and adherence to intervention:* Adherence will be assessed by the completion of sessions, and the use of the goal setting and health tracking features. We will also assess patterns and frequency of programme usage.
- *Attrition rate:* The number of dropouts (attrition rate) will be recorded.
- *Missing data:* Quantity of missing data (e.g., questionnaire return rate, outcome measures not completed)
- *Patient experience:* This will be attained through qualitative interviews in a sub-set of patients.

#### Primary study outcome

The primary outcome is the Patient Activation Measure (PAM-13), the most widely used instrument for measuring patient activation ^8^ which was piloted in the NHS through the UK Renal Registry (UKRR), and has been validated ^23^ and recommend for use in those with kidney disease ^14 23^. The PAM-13 is a validated tool of 13 questions which assesses a patient’s knowledge, skills, and confidence in managing their own health. The PAM-13 has demonstrated good internal consistency as well as adequate reliability and validity ^8 24^, including in those with kidney disease ^23^. Answers are weighted and combined to provide a score on a scale from 0 to 100. The PAM allows respondents to be categorized into one of four levels with lower levels indicating low activation and higher levels indicating high activation. The PAM-13 will be assessed amongst a battery survey of other questionnaires that will be delivered online using Jisc Online Surveys (University of Leicester). As per SPIRIT recommendations, the timepoints for each outcome can be found in **Table 2**.

**Table 2.**
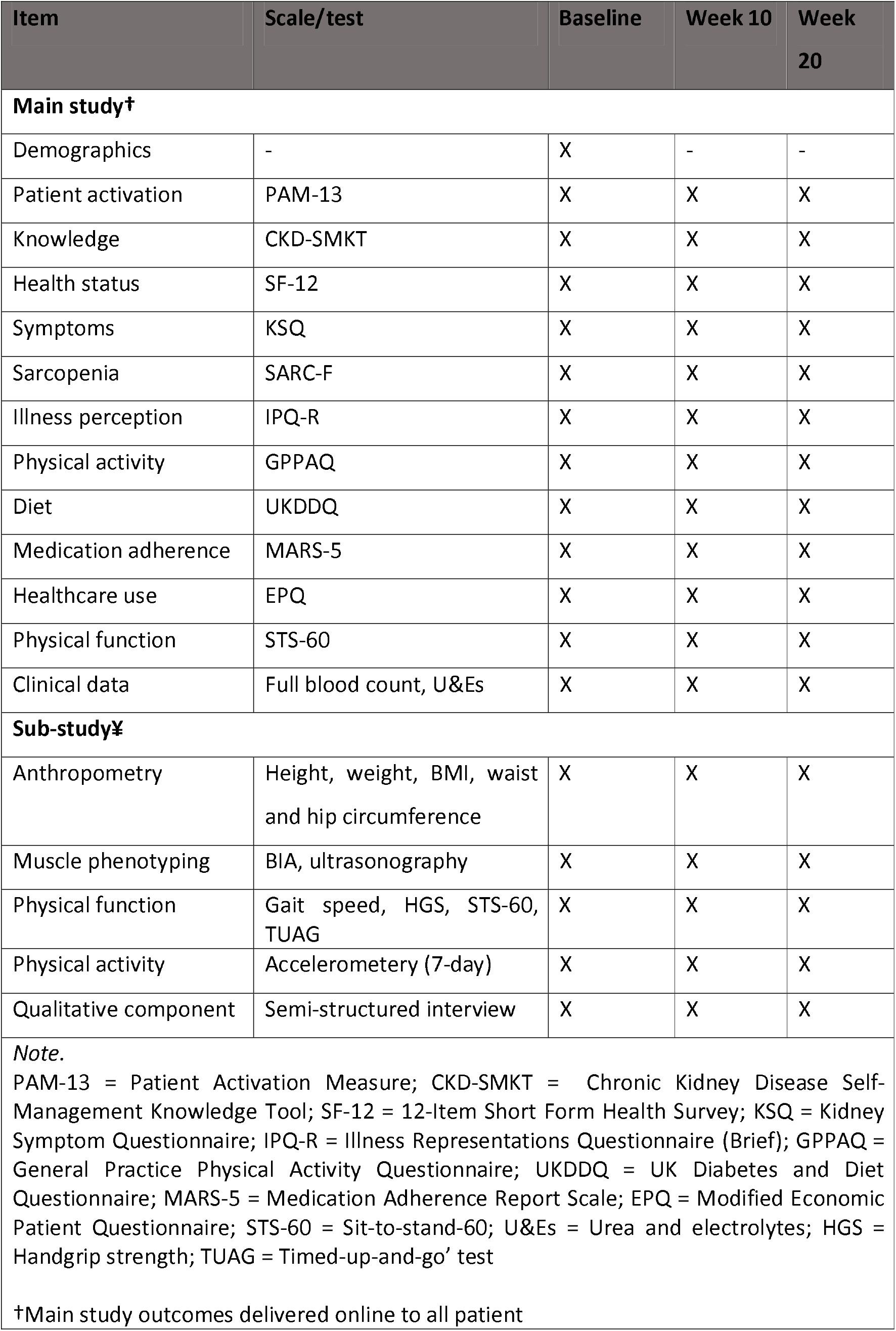
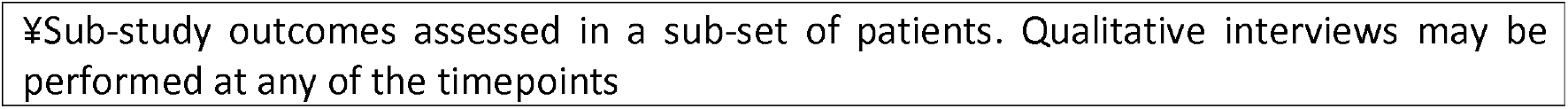
Outcome measure timepoints.

| Item | Scale/test | Baseline | Week 10 | Week 20 |
| --- | --- | --- | --- | --- |
| **Main study†** |  |  |  |  |
| Demographics | - | X | - | - |
| Patient activation | PAM-13 | X | X | X |
| Knowledge | CKD-SMKT | X | X | X |
| Health status | SF-12 | X | X | X |
| Symptoms | KSQ | X | X | X |
| Sarcopenia | SARC-F | X | X | X |
| Illness perception | IPQ-R | X | X | X |
| Physical activity | GPPAQ | X | X | X |
| Diet | UKDDQ | X | X | X |
| Medication adherence | MARS-5 | X | X | X |
| Healthcare use | EPQ | X | X | X |
| Physical function | STS-60 | X | X | X |
| Clinical data | Full blood count, U&Es | X | X | X |
| **Sub-study‡** |  |  |  |  |
| Anthropometry | Height, weight, BMI, waist and hip circumference | X | X | X |
| Muscle phenotyping | BIA, ultrasonography | X | X | X |
| Physical function | Gait speed, HGS, STS-60, TUAG | X | X | X |
| Physical activity | Accelerometry (7-day) | X | X | X |
| Qualitative component | Semi-structured interview | X | X | X |
| *Note.*<br>PAM-13 = Patient Activation Measure; CKD-SMKT = Chronic Kidney Disease Self-Management Knowledge Tool; SF-12 = 12-Item Short Form Health Survey; KSQ = Kidney Symptom Questionnaire; IPQ-R = Illness Representations Questionnaire (Brief); GPPAQ = General Practice Physical Activity Questionnaire; UKDDQ = UK Diabetes and Diet Questionnaire; MARS-5 = Medication Adherence Report Scale; EPQ = Modified Economic Patient Questionnaire; STS-60 = Sit-to-stand-60; U&Es = Urea and electrolytes; HGS = Handgrip strength; TUAG = Timed-up-and-go' test<br>†Main study outcomes delivered online to all patient |  |  |  |  |

¥Sub-study outcomes assessed in a sub-set of patients. Qualitative interviews may be performed at any of the timepoints

#### Secondary outcomes

Alongside a bespoke self-reported demographic questionnaire, other validated questionnaires in the online survey, which capture different aspects of self-management and lifestyle behaviours that are addressed in ‘My Kidneys & Me’, include:

- *Chronic Kidney Disease Self-Management Knowledge Tool (CKD-SMKT):* The CKD-SMKT is a validated 11-item questionnaire, which assesses kidney disease patients’ knowledge of various key self-management behaviours and kidney health ^25^.
- *12-Item Short Form Health Survey (SF-12):* The SF-12 is a multipurpose short form survey with 12 questions. The SF-12 assesses patients mental and physical functioning and overall health-related-quality of life (QoL) ^26^.
- *Kidney Symptom Questionnaire (KSQ):* The KSQ assesses the frequency, intrusiveness, and total impact of a range of 13 common kidney disease-related symptoms. This questionnaire has been validated by our group ^27^ and used widely in the literature ^28^.
- *SARC-F questionnaire:* The SARC-F questionnaire includes five components: strength, assistance walking, rise from a chair, climb stairs, and falls. SARC-F scores range from 0 to 10 and a score ≥4 is predictive of sarcopenia ^29^.
- *Illness Representations Questionnaire (Brief) (IPQ-R):* The IPQ-R is a widely accepted measure of illness representations. These components are identity, cause, timeline, consequence, and controllability/cure. The questionnaire consists of three parts: an identity scale, a structure scale, and a causal scale ^30^.
- *General Practice Physical Activity Questionnaire (GPPAQ):* The GPPAQ was developed by the World Health Organization and Department of Health detailing a 4-level Physical Activity Index (PAI) reflecting an individual’s current physical activity. The GPPAQ is a validated measure of physical activity behaviour in CKD ^31^.
- *UK Diabetes and Diet Questionnaire (UKDDQ):* The UKDDQ is a 25-item questionnaire designed to assess diet and dietary behaviours. Respondents are asked to identify how often they consumed certain foods (vegetables, fruits, sugary drinks, processed meat) over that last month ^32^. Responses will be scored as per previous research ^33^. This questionnaire is sensitive to changes following an intervention ^34^.
- *Medication Adherence Report Scale (MARS-5):* The MARS-5 is a non-disease specific questionnaire used to measure adherence to medications and has been previously used in CKD. This comprises of five questions regarding changing medication dosage, forgetting to take medication, consciously stopping taking medication, skipping medication, and using less than prescribed ^35^.
- *Modified Economic Patient Questionnaire (EPQ):* We will use a modified version of the ‘Economic Patient Questionnaire (EPQ)’ to assess participants’ use of inpatient and outpatient services and data on non-hospital-based health and social care use at all assessment points ^36^.

When the participant consents to the study, the researcher will access their clinical records and extract information such as blood and urine results to gather information of kidney function, proteinuria, iron status, and lipid profiles. We will record prescribed medication. Self-reported co-morbidities will be checked against those recorded in the medical notes.

#### Sub-study

In an optional sub-study, the following additional physical assessments will be performed during a visit to a hospital site:

- *Anthropometry:* Height, weight, body mass index (BMI), and waist and hip circumference will be measured in line with established procedures. Resting heart rate and blood pressure will be assessed using a standard sphygmomanometer device.
- *Muscle phenotyping:* Muscle and fat mass/size/thickness, and body fat % will be measured using a free-standing bioelectrical impendence analysis (BIA) monitor and B-mode ultrasonography of the rectus femoris muscle. These are painless, non-invasive methods for measuring body composition. Our group has recently validated our BIA device against dual-energy X-ray absorptiometry (DXA) ^37^ whilst ultrasonography can be used to assess sarcopenia in CKD ^38 39^.
- *Gait speed:* The participant is asked to walk a 4m course at their ‘usual’ walking speed, with a walking aid if normally used ^40^.
- *Handgrip strength (HGS):* Handgrip strength will be assessed using a handheld dynamometer (Jamar Plus+ Digital). Participants will be asked to hold the dynamometer with their shoulder at 0 degrees flexion and elbow at 90-degree flexion. Participants will be asked to squeeze the dynamometer handle maximally for 3 seconds. The peak force output (kg) from the three attempts from both the dominant and non-dominant hand will be recorded ^41^.
- *Sit-to-stand-60 test:* Sit-to-stand (STS) tests are a good measure of functional ability and have been used extensively in CKD patients. Our group has extensive use and knowledge of this test, and published reliability and validation data for it ^42^. The patient starts from a seated position on a hard, upright chair, with the feet flat on the floor and the knees bent at 90°. For the test, the patient simply stands up fully and then sits down again to the starting position, without using the hands (one repetition). The STS-60 test involves completing as many STS cycles as possible in 60 seconds. Participants will also be asked to record a STS-60 score as part of the online survey.
- *Timed-up-and-go’ test (TUAG):* The TUAG assesses mobility and requires dynamic balance. The participant will be asked to rise from a chair, walk 3 metres, turn around a cone, and sit back down ^40^.
- *Accelerometery:* To accurately measure objective physical activity, patients will wear a wrist accelerometer (GENEActiv) for a 7-day period before the intervention and post-intervention ^31 43^.

Familiarisation of objective physical performance measures will be performed before baseline assessments. In order to assess any potential differences in administering the questionnaires online, participants in the sub-study will also be asked to complete the same questionnaires via paper format.

In the sub-study, semi-structured interviews will be held with a researcher trained in qualitative methodology. Individual interviews will last between ∼30 to 60 minutes. Interviews will take place in private area. For those unable to attend a face-to-face interview, interviews may also be performed via telephone using a secure recording device as used currently in other studies by our group. Patients recruited may be interviewed on at least one occasion (e.g., before and/or after the intervention). Topics in this interview will include current self-management knowledge, skills, confidence and behaviours, and attitudes towards lifestyle self-management. Topics of the subsequent interviews will include experiences of the intervention, the quality of the content, the quality of the delivery method, reasons for non-adherence with the intervention, and healthcare usage.

### Patient and public involvement

A full description of how patients and their families were involved in the development of the programme can be found in Lightfoot et al. ^21^. In summary, a study patient steering group was formed consisting of ten individuals living with kidney disease and two family members. An initial priority setting workshop determined key topics of interest to this group, including lack of educational support from healthcare professionals. Following this, we developed an educational booklet which was co-designed using our patient steering group. This booklet was disseminated to local primary care practices where individuals provided feedback on the content. The information in this booklet was then adapted for digital use and was termed ‘My Kidneys & Me’ by the patient group. We also performed semi-structured interviews with patients around self-management and the use of educational resources. Key examples of feedback from patients included the following: 1) to provide a symptom tracker to enable self-monitoring of symptoms; 2) include a separate session on sleep; 3) include a separate session on well-being, emphasising the importance of looking after mental health; 4) use of myth and fact quizzes as a way to test knowledge; 5) state how long sessions should take in the introduction. Once ‘My Kidneys & Me’ was developed, members of the steering group were provided with access to the programme to provide initial comment. The patient steering group assisted with the selection of questionnaires for the study, and reviewed the final questionnaire survey to ensure that it was acceptable. Throughout the study, the steering group will be used to develop a suitable and relevant topic guide exploring exploration of their attitudes towards self-management and the impact these have on their lives. The group will also be used to interpret initial and final findings. In addition, the patient steering group will support the development of the lay summary outputs to be disseminated to patients and the public.

### Data analysis

#### Sample size calculation

With PAM-13 as the primary outcome in the full RCT, a total sample size of n=432 participants are required based on a 2:1 randomisation (n=288 in the intervention group and n=144 in the control group). This was based on previously published PAM-13 data by our group ^13^ and on the required power to detect a minimal clinically significant difference of 4 points in the PAM-13 ^44-46^. For the initial nested pilot study, a pragmatic sample size of n=60 participants (n=40 in the intervention group and n=20 in the control group, based on 2:1 randomisation) will be used. We aim to recruit at least 10 patients into the optional sub-study.

#### Statistical analysis

Participant demographics and clinical characteristics will be analysed using descriptive statistics. The primary outcome is the PAM-13 where answers are weighted provide a score on a scale from 0 to 100. As recommended by Twisk et al. ^47^, estimates of treatment effect will be assessed by longitudinal analysis of covariance. In this method the outcome variable measured at the different follow-up measurements (post-intervention at week 10, follow-up at week 20) is adjusted for the baseline value of the outcome. Analysis will be performed using ‘intention-to-treat’ (ITT) analysis. In an ITT approach, patients are analyzed by how they were randomised regardless of their actual compliance with treatment. For patient-reported outcomes missing data items will be handled according to established protocols for the validated surveys. Additional post-hoc analysis will be performed to determine if differences exist in PAM score changes between those with low and high PAM scores.

In the sub-study, interview recordings will be professionally transcribed verbatim. The precise analytical methodology used may change based on the nature of the data collected, however thematic analysis will be used as an initial foundation for data analysis as it provides a systematic model for managing and mapping the data. Analysis will follow recognised steps (e.g., familiarisation, initial and confirmation of coding (using NVivo), defining themes). In an integrative strategy, any available quantitative data will be used to inform the qualitative analysis.

## Data Availability

All data produced in the present study are available upon reasonable request to the authors

## ETHICS AND DISSEMINATION

To carry out this study, we will consider the Good Clinical Practice (GCP) guidelines of the local governance organisations (University of Leicester and University Hospitals of Leicester NHS Trust) thus guaranteeing the protection of the rights, the safety, and the well-being of the participants of the trial in compliance with the principles of the Declaration of Helsinki, as well as the credibility of the data obtained in the clinical trial. All participant data entered into the ‘My Kidneys & Me’ will be managed through the MyDESMOND platform. Data entered as part of the online outcomes is managed by Jisc Online Surveys under a University of Leicester license. A full informative sheet explaining the study in detail, the voluntary nature of the research, and the procedure for the protection of their personal data will be provided along with the online consent form. Patients will be given the opportunity to contact the researchers with any questions prior to the informed consent. The informed consent obtained from study participants will be online.

This study was fully approved by the Research Ethics Committee (REC)-Leicester South and Health Research Authority (HRA) on the 19/11/2020 (reference: 17/EM/0357). The results are expected to be published in scientific journals and presented at clinical research conferences in 2024. Findings will be disseminated to patients and the wider kidney and healthcare community via social media platforms, interest groups, recruiting sites, and institutions associated with the research team. Any subsequent changes to the study protocol will reviewed by the REC and HRA through appropriate amendments. Any significant protocol changes will be stated on the ISRCTN Registry. The study was prospectively registered as ISRCTN18314195.

## SUMMARY

‘My Kidneys & Me’, an online self-management and lifestyle education programme, aims to increase patient activation and promote self-management behaviours. If this is achieved, it is anticipated that the intervention will achieve its distal aims of improving patient QoL, physical function, and symptom burden whilst experiencing fewer hospital admissions and saving costs to the NHS.

## LIST OF ABBREVIATIONS

BIA: Bioelectrical Impendence Analysis
BMI: Body Mass Index
CKD: Chronic Kidney Disease
CKD-SMKT: Chronic Kidney Disease Self-Management Knowledge Tool
COVID-19: Coronavirus Disease-2019
DXA: Dual-Energy X-Ray Absorptiometry
eGFR: Estimated Glomerular Filtration Rate
EPQ: Modified Economic Patient Questionnaire
EWGSOP: European Working Group on Sarcopenia in Older People
GCP: Good Clinical Practice
GP: General Practitioner
GPPAQ: General Practice Physical Activity Questionnaire
HGS: Handgrip Strength
HRA: Health Research Authority
IPQ-R: Illness Representations Questionnaire
ITT: Intention-To-Treat
KSQ: Kidney Symptom Questionnaire
MARS-5: Medication Adherence Report Scale
NHS: National Health Service
NICE: National Institute for Health and Care Excellence
PAI: Physical Activity Index
PAM-13: Patient Activation Measure
QoL: Quality of Life
RCT: Randomised Control Trial
REC: Research Ethics Committee
SF-12: 12-Item Short Form Health Survey
SPIRIT: Standard Protocol Items: Recommendations for Interventional Trials
STS: Sit-To-Stand
TUAG: Timed-Up-And-Go
UKDDQ: UK Diabetes and Diet Questionnaire
UKRR: UK Renal Registry

## DECLARATIONS

### Ethics approval and consent to participate

This study was fully approved by the Research Ethics Committee-Leicester South on the 19/11/2020 (reference: 17/EM/0357). All participants will provide consent either online or face-to-face (sub-study). All participants will be given the opportunity to ask questions before completing the consent process. The trial is sponsored by the University of Leicester.

### Consent for publication

N/A

### Availability of data and materials

N/A

### Authors’ contributions

All authors contributed to the design of the SMILE-K trial protocol. CLJ and TJW drafted the manuscript contributing equally. All authors read and approved the final manuscript.

## Acknowledgements

The authors would like to thank participating members of our patient and public steering group in their support in the development of the study. The authors thank members of the professional stakeholder group who have contributed to the design and development of ‘My Kidneys & Me’: Jonathan Barratt, Mike Bonar, Christopher Brough, James Burton, John Feehally, Charlie Franklin, Matthew Graham-Brown, Michelle Hadjiconstantinou, Jenny Hainsworth, Vicki Johnson, Maria Martinez, Andrew Nixon, Vicky Pursey, Sally Schreder, Hannah Young, Noemi Vadazsy, Fiona Willingham, and Lucina Wilde.

